# Epigenetic regulation of inflammation in post-operative organ dysfunction: a scoping review protocol

**DOI:** 10.1101/2025.03.03.25323001

**Authors:** Ruairí Wilson, Charlotte Fern, Carl Goodyear, Ben Shelley

**Affiliations:** Anaesthesia, Perioperative Medicine and Critical Care Research Group, School of Medicine, University of Glasgow; School of Infection and Immunity, University of Glasgow; Department of Anaesthesia, Golden Jubilee National Hospital, Clydebank

## Abstract

**Introduction:** The inflammatory response to surgery is complex, dynamic and exhibits variability in magnitude and duration among patients undergoing similar operations. Dysregulated inflammation is associated with post-operative organ dysfunction, particularly after major surgery. Epigenetic modifications enable (or prohibit) selective gene transcription without altering DNA sequences, effectively regulating gene expression. Several studies have investigated epigenetic regulation of the immune system in the context of surgery, often studying organ-specific dysfunction.

**Objectives:** We propose a novel scoping review protocol to collate and synthesise existing studies investigating epigenetic regulation of post-operative inflammation, as a key mechanism of post-operative organ dysfunction and complications. We will map knowledge gaps to inform future research in this emerging field.

**Methods and analysis:** This scoping review protocol has been created following the Joanna Brigg’s Institute (JBI) updated guidelines for conducting scoping reviews. The protocol has been further examined alongside the Preferred Reporting Items for Systematic reviews and Meta-Analyses (PRISMA) extension for Scoping Reviews (PRISMA-ScR) checklist and is registered on Open Science Framework (doi.org/10.17605/OSF.IO/CE8FB). Published human studies from 1946 to the present will be considered. Studies will include patients undergoing surgery, where epigenetic regulation of the immune system is investigated alongside assessment of organ dysfunction or complications. Searches will be conducted using Medline (via OVID) and Embase. Two reviewers will independently screen titles, abstracts and full texts of studies meeting the inclusion criteria. Following study screening, a customised data extraction form will collect study information related to the review questions and inclusion criteria (population, concept, context). Results will be presented by diagrammatic mapping of studies and tabular representation of findings.

## Introduction

Post-operative complications are a major cause of morbidity, mortality and increased healthcare cost in patients undergoing surgery[1-3]. The ‘stress response’ to surgical trauma results from well-choreographed activation of autonomic, neuroendocrine, metabolic, and inflammatory responses, necessary to maintain host homeostasis and facilitate tissue repair. Unchecked inflammation can however have detrimental effects on capillary permeability, immune function, wound healing and organ function, culminating in post-operative complications. Serological markers of inflammation have identified that the magnitude of the post-operative inflammatory response is associated with post-operative complications following surgery[4-6]. The degree of post-operative inflammation is associated with multi-organ morbidities, including: myocardial injury[7], lung injury[8], post-operative cognitive dysfunction[9] and acute kidney injury[10].

A ‘genomic storm’ occurs within hours of major surgery, affecting more than 80% of cellular pathways, dynamically altering the leucocyte transcriptome, with upregulation of the innate immune system and suppression of adaptive immune responses [3, 11]. In patients undergoing major thoracoabdominal surgery, Allen et al demonstrated differential expression of 522 genes governing leukocyte function 24-hours after surgery; 248 (48%) associated with innate inflammation were upregulated while 274 (52%) associated with adaptive immunity were downregulated[11]. In this study, within-patient gene expression change was associated with postoperative infection, hospital length of stay, and outcomes were worse in patients in whom postoperative gene expression was most radically altered.

The potential for host genomics to impact peri-operative outcomes has received some study, and whilst it has been possible to identify single nucleotide polymorphisms which might lead to specific perioperative morbidity syndromes (e.g. butyrylcholinesterase deficiency[12]), it is unlikely that polymorphisms are responsible for the wide heterogeneity seen in responses to surgery. The value of studying genetic susceptibility to complications is further limited by the static nature of the genome; should predictors of outcome be identified, these genetic changes are fixed, with limited potential for therapeutic intervention. By contrast, the epigenome is dynamic and amenable to therapeutic manipulation or upstream modification. Future epigenetic study therefore has potential to predict and modify perioperative risk.

### Epigenomics and inflammation

Epigenomics concerns the study of how cells control gene activity without changing the DNA sequence. Cellular DNA is bound in chromatin as a complex of DNA and proteins. Modification of these proteins (classically by DNA methylation or histone modification) enables differential gene expression, changing the way a cell responds to a stimulus. Classical descriptions of the immune system describe the innate and adaptive responses, with only the adaptive immune response able to build immunological memory. In recent years, epigenetic reprogramming of innate immune cells in response to stimulation has been recognised [13, 14]. Following an initial challenge from an inflammatory insult, cells return to a non-activated state, but through modification to the epigenome, are primed for response to a subsequent stimulus. After a secondary challenge, rapid and enhanced recruitment of transcription factors escalate the pro-inflammatory phenotype.

Epigenetic modifications have been extensively studied in cancer, particularly as prognostic markers [15, 16]. Comparatively few studies investigate epigenetic involvement in mediating post-operative organ dysfunction. Initial searches in this area have identified a small number of prospective observational studies. Several of these studies investigate epigenetic mechanisms of post-operative delirium [17-21], most commonly in the context of neurosurgery [18-21]. Epigenetic modification mediated through DNA methylation has also been associated with increased chronic post-surgical pain in a large prospective cohort of adolescents undergoing spinal surgery for scoliosis [22]. In patients undergoing major cardiac surgery, Laudanski and colleagues initially demonstrated epigenetic regulation of pro-inflammatory gene activity in monocytes of 20 patients [23], but did not observe significant organ dysfunction. This group then conducted a larger study of 59 patients undergoing cardiac surgery, demonstrating prolonged serological histone release and inflammation, persisting 3-months after surgery [24]. Fischer et al have demonstrated epigenetic susceptibility to atrial fibrillation following cardiac surgery [25, 26], a common complication in this patient cohort. In an elderly population undergoing major colorectal and orthopaedic surgery, Sadahiro and colleagues reported DNA methylation as a key mechanism influencing inflammation after surgery, describing rapid post-operative methylation, with stable expression 7-days post-operatively [27].

This scoping review offers a novel and timely opportunity to collate and map studies investigating organ-specific complications of inflammatory epigenetic regulation in the context of surgery and to identify the smaller number of studies which assess complications in multiple organ systems and in different surgical contexts. Mapping available evidence will inform current understanding, helping to guide future research design in this expanding field.

### Aim

To identify and map existing evidence of inflammatory post-operative organ dysfunction (and complications) mediated through epigenetic regulation.

In keeping with the Joanna Brigg’s Institute (JBI) updated guidelines for scoping reviews [28-30], we will systematically:

- identify types of evidence available;
- identify and analyse knowledge gaps;
- determine key concepts and definitions (for example, how epigenetic modifications are defined and described in the literature);
- examine how research is conducted;
- identify key factors related to epigenetic-regulated inflammation in the context of surgery.

### Review Questions

Primary question

1. What evidence exists for epigenetic-regulated inflammation as a mechanism of organ dysfunction after surgery?

Secondary questions

2a: What types of studies are available?

2b: How are epigenetic modifications defined and described in the literature?

2c: What specific organ dysfunctions (or other complications of surgery) have been studied?

2d: What types of surgery have been studied?

2e: Where are the knowledge gaps in the field?

## Methods

This scoping review protocol has been created following guidance from the updated JBI guidelines for conducting scoping reviews [28, 29] and the Arskey and O’Malley framework [31]. The protocol has been further examined alongside a completed Preferred Reporting Items for Systematic reviews and Meta-Analyses (PRISMA) Extension for Scoping Reviews (PRISMA-ScR) checklist [32] (supplementary appendix 1). A scoping review is appropriate for our exploratory review questions, which aim to systematically identify, collate and map available evidence pertinent to our research questions and inclusion criteria. We additionally aim to identify gaps in knowledge to guide future research focus.

### Inclusion criteria

This scoping review will include all available human studies which investigate organ dysfunction (or complications) resulting from epigenetic mechanisms of inflammation in the context of surgery (table 1).

**Table 1:**
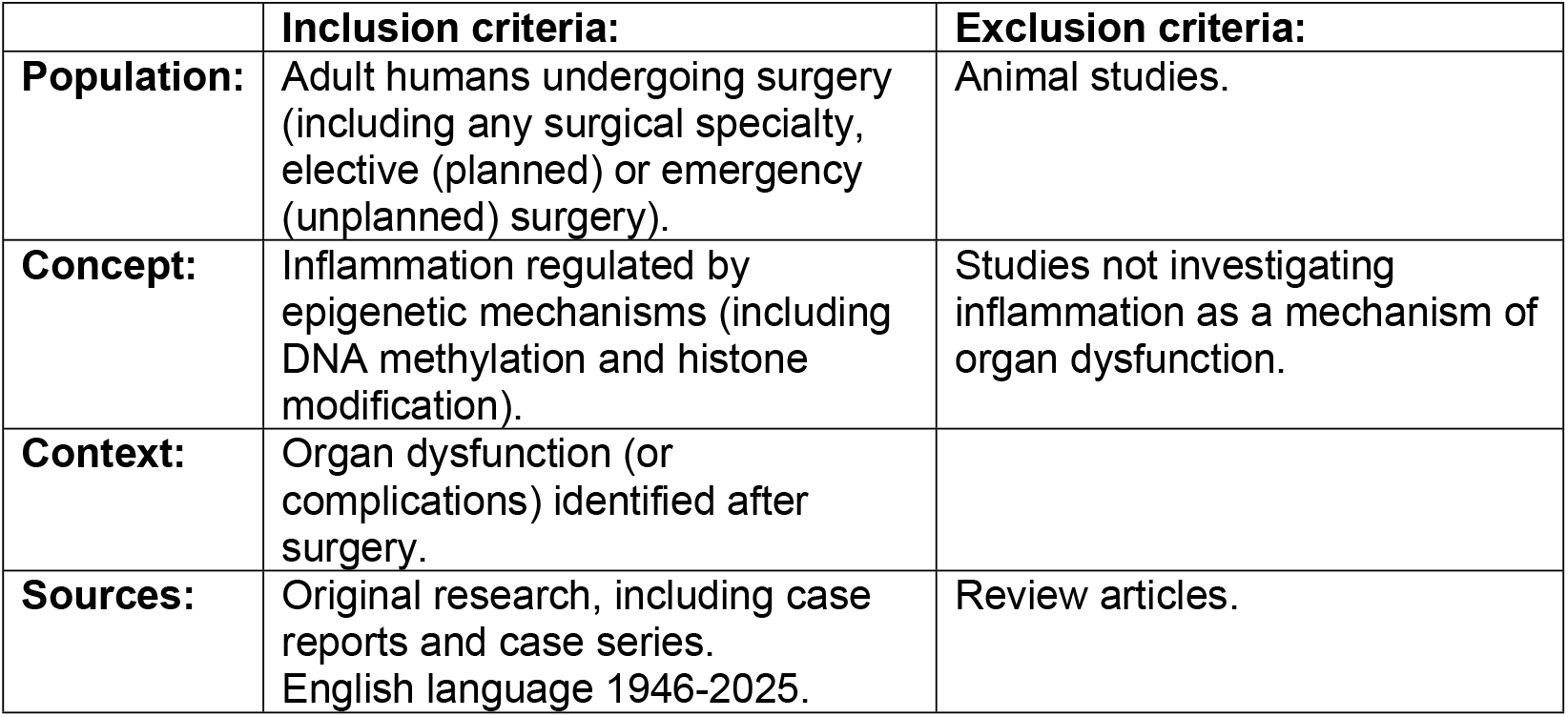
Inclusion and exclusion criteria. formatted by JBI-recommended Population, Concept, Context (PCC) framework.

### Population

Humans undergoing any type of surgery, including unplanned emergency or planned elective operations.

### Concept

Inflammation regulated by epigenetic mechanisms. Epigenetic modification typically occurs through DNA methylation, or histone modification. One sub-question for this review (2b) seeks to address the heterogeneity of terms used to describe epigenetic modifications.

### Context

Organ dysfunction (or complications) identified after surgery. We anticipate several studies will focus investigation on a particular organ dysfunction (for example, post-operative delirium) while other studies will assess for a range of possible post-operative complications. This review aims to collate and map all available evidence regarding post-operative organ dysfunction (or complications) mediated through epigenetic regulation of inflammation, and we will therefore include all studies concerning post-operative complications mediated through this mechanism.

### Exclusion criteria

Animal studies will be excluded owing to concerns of translatability and relevance to human organ dysfunction in the context of surgery. Review articles will be excluded given the need to collate and map original research.

### Databases

Searches will be conducted in Medline (via OVID) and Embase.

### Search strategy

The search strategy aims to locate published studies using the JBI three-step approach [26, 27]. An initial pilot search of Medline (via OVID) and Embase will be undertaken to identify relevant articles. Keywords and index terms used in the title and abstracts of relevant papers will be used to generate a full search strategy for Medline (such as the pilot search in supplementary appendix 2) and Embase. The search strategy will be adapted for Embase. The reference list of all included studies will be screened for additional studies. Studies published in English from 1946 to present day will be included for screening. The citation manager software Endnote 21.4 will be used to remove duplicate articles and the online software tool Covidence (Covidence systematic review software, Veritas Health Innovation, Melbourne, Australia) will be used to apply inclusion and exclusion criteria (table 1) for selection of articles during screening.

### Study selection

At least two independent reviewers will conduct literature screening of titles and abstracts using the inclusion and exclusion criteria. Selected articles will undergo secondary screening of full-text articles prior to data extraction (figure 1). Reviewers will meet regularly to resolve disagreements through discussion of the article in question against the inclusion and exclusion criteria. Where discussion cannot resolve disagreement, an additional reviewer will assist until consensus is reached. The systematic literature search will be presented in the format of a PRISMA-ScR flow chart for reproducibility and transparency [29]. Results will be reported using descriptive statistics, diagrammatic figures or tables of information, in addition to narrative descriptions in keeping with the JBI guidelines [27].

**Figure 1:**
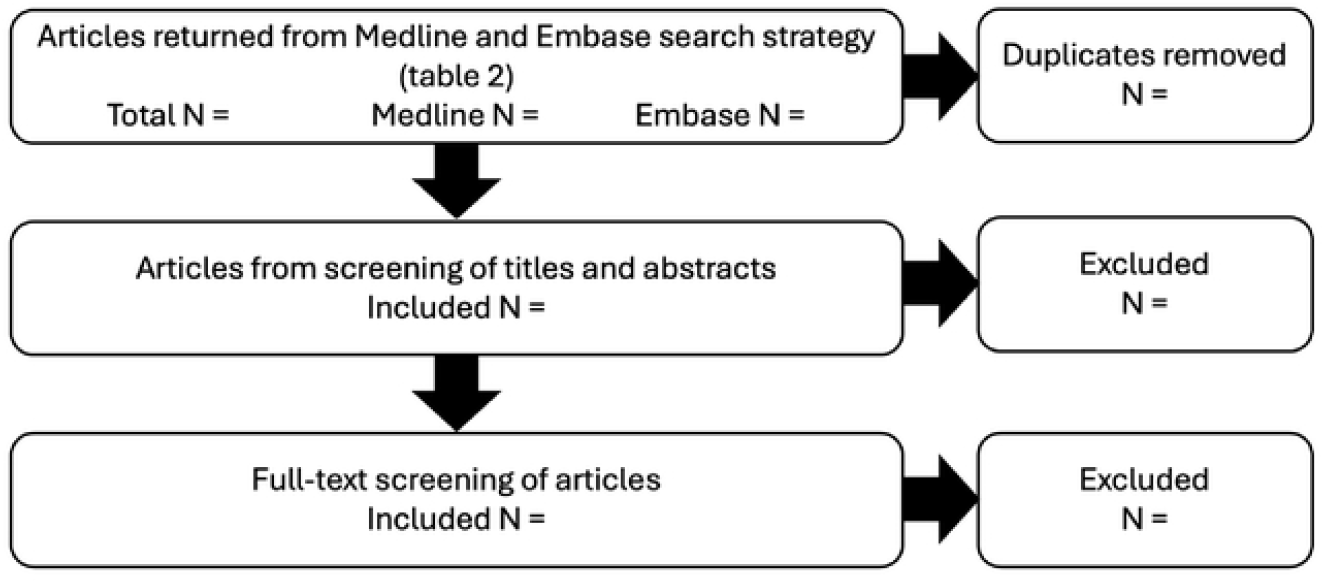
Flow chart showing study selection procedure.

### Data extraction

Data extraction will similarly be performed by at least two independent reviewers, followed by cross-checking of included articles. Disagreements will be resolved through discussion and mutual agreement of the article in question. Where this fails to resolve a disagreement, consultation with an additional independent reviewer will be undertaken until mutual agreement. Data extraction will include basic descriptive data of each article and information relating to the review question and inclusion criteria. A draft table detailing anticipated data extraction is shown in table 2, and includes population, concept and context subheadings of the inclusion criteria, as recommended by the JBI [26, 27]. The draft data extraction tool will be piloted on 20% of the articles screened for inclusion to test for feasibility and utility in answering the review questions. We anticipate that the data extraction tool may be amended during the review process to enhance our ability to answer the research questions. Disclosure and justification for modifications will be detailed in the subsequent scoping review report.

**Table 2:**
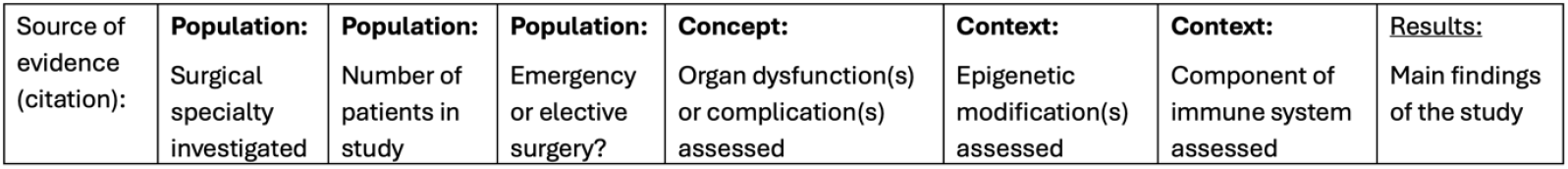
Anticipated data extraction table, including columns for basic descriptive data, population-, concept- and context-specific study features related to the research questions and inclusion criteria.

| Source of evidence (citation): | Population: | Population: | Population: | Concept: | Context: | Context: | Results: |
| --- | --- | --- | --- | --- | --- | --- | --- |
|  | Surgical specialty investigated | Number of patients in study | Emergency or elective surgery? | Organ dysfunction(s) or complication(s) assessed | Epigenetic modification(s) assessed | Component of immune system assessed | Main findings of the study |

### Data analysis

Descriptive data analyses including frequency analysis of studies will be presented. We anticipate this will involve diagrammatic mapping of studies and tabular representation of findings. Diagrammatic representation is likely to be of particular importance to demonstrate the different types of surgery investigated, organ dysfunctions investigated, and the population sizes recruited to each study. Quality assessment of studies will not be performed as this is beyond the remit of a scoping review, our primary objective is to collate and comprehensively map available evidence.

### Protocol deviations

We aim to conduct our scoping review process in-line with the principles and steps outlined, in keeping with best-practice guidelines [26-29]. However, given the iterative nature of the scoping review process, protocol deviation may be necessary during the search strategy, data extraction or data presentation stages. Disclosure and justification for deviations to the described protocol will be detailed in the subsequent scoping review report.

## Conclusions

This scoping review proposes a novel approach to collate existing studies on an important and expanding research area. We aim to map existing studies, to identify knowledge gaps which will inform future research design. The strengths of this study include the novel design, inclusion of diverse study types (different types of surgery, assessing a range of organ-specific and systemic complications of surgery) and the mapping of knowledge gaps. Limitations include restriction of included articles to English language.

## Supporting information

Supplementary appendix 2: MEDLINE pilot search

## Data Availability

All relevant data from this study will be made available upon study completion.

## Data availability

No original data sets will be generated in this proposed study. Relevant data extracted from synthesis of available study evidence will be detailed as specified in the subsequent scoping review report.

## Funding statement

No specific sources of funding were acquired for this work. This work is supported by the sponsoring institution, the University of Glasgow. BS is supported by the National Institute of Academic Anaesthesia / Royal College of Anaesthetists, British Oxygen Company Chair grant.

## Acknowledgement

The authors wish to thank Paul Cannon, Librarian at the University of Glasgow College of Medicine, Veterinary and Life Sciences for his contribution and advice in preparation of the search strategy.

## Notes

### Competing Interest Statement

The authors have declared no competing interest.

### Funding Statement

The author(s) received no specific funding for this work.

