## Supplementary appendix 2: MEDLINE pilot search for "Epigenetic regulation of inflammation in post-operative organ dysfunction: a scoping review protocol"

| **Database:** | **Search strategy:** |
| --- | --- |
| Medline (via OVID) | Ovid MEDLINE(R) ALL <1946 to February 20, 2025>  **Line: Search term: Results:**  1 surgery/ 41334  2 anaesthesia/ 225  3 anesthesia/ 67950  4 anesthesiology/ 34097  5 postop*.mp. 1072714  6 post-op*.mp. 108269  7 after surgery.mp. 210122  8 surgical.mp. 1702084  9 epigen*.mp. 149196  10 dna meth*.mp. 99282  11 histone mod*.mp. 18333  12 histon*.mp. 149276  13 h3k4*.mp. 7343  14 h3k27*.mp. 9945  15 genetic.mp. 2018565  16 or/1-8 2522836  17 or/9-15 2194577  18 organ d*sfunction.mp. 19974  19 organ injur*.mp. 7379  20 organ failure.mp. 36818  21 complicat*.mp. 3911480  22 postoperative complication*.mp. 477698  23 adverse event*.mp. 262097  24 adverse effect*.mp. 2264274  25 morbidity.mp. 508514  26 mortality.mp. 1531217  27 respiratory failure.mp. 45368  28 hypox*.mp. 253315  29 acute respiratory distress syndrome.mp. 25726  30 pneumonia.mp. 242468  31 shock.mp. 276788  32 hypotension.mp. 79611  33 arrhyth*.mp. 177660  34 myocard*.mp. 658015  35 ischaem*.mp. 71363  36 ischem*.mp. 481465  37 lactate.mp. 157272  38 cardiac death.mp. 33276  39 cardiac arrest.mp. 49538  40 pulmonary embolism.mp. 65559  41 deep venous thrombosis.mp. 13834  42 atrial fibrillation.mp. 117962  43 coma.mp. 53822  44 delirium.mp. 27013  45 confus*.mp. 77883  46 stroke.mp. 412212  47 cerebrovascular accident.mp. 5990  48 pain.mp. 955100  49 kidney injury.mp. 87567  50 kidney failure.mp. 114900  51 renal injury.mp. 15098  52 renal failure.mp. 100119  53 kidney dys*.mp. 4310  54 renal dys*.mp. 24460  55 acute kidney injury.mp. 79666  56 anaem*.mp. 41906  57 anem*.mp. 224862  58 neutro*.mp. 309218  59 leuko*.mp. 429833  60 lympho*.mp. 1191142  61 thrombo*.mp. 550019  62 platel*.mp. 335682  63 pancyto*.mp. 11716  64 marrow suppres*.mp. 2891  65 myelo*.mp. 391735  66 coagulop*.mp. 21339  67 disseminated intravascular coag*.mp. 18619  68 liver failure.mp. 33228  69 liver dys*.mp. 10791  70 hepatic fail*.mp. 9976  71 hepatic dys*.mp. 5625  72 fever.mp. 273072  73 febrile.mp. 45268  74 infect*.mp. 2944243  75 systemic inflammatory response syndrome.mp. 11971  76 sequential organ failure assessment.mp. 6187  77 apache.mp. 15031  78 multiple organ dysfunction score.mp. 202  79 clavien-dindo.mp. 7999  80 surgical site infection.mp. 13614  81 critical care.mp. 96626  82 critical illness.mp. 50135  83 inflam*.mp. 1503303  84 immun*.mp. 4199554  85 c-reactive protein.mp. 113179  86 interleukin*.mp. 440323  87 tumor necrosis factor.mp. 239937  88 exp human/ 22540147  89 review/ 3435834  90 or/18-82 12466398  91 or/83-87 5272693  92 (16 and 17 and 88 and 90 and 91) not 89 1995  93 limit 92 to english language 1863 |

**Supplementary appendix 2: Table showing a pilot search strategy for the Medline (via OVID) database.**
